# Real-world effectiveness of antipsychotic treatment of functional outcomes over 10 years: A national cohort of patients in Denmark with schizophrenia

**DOI:** 10.64898/2026.01.07.26343585

**Authors:** Ricardo Twumasi, Frederikke Hørdam Gronemann, Carsten Hjorthøj, Oliver D.Howes, Maximin Lange, Merete Nordentoft, Merete Osler

## Abstract

**Background:** Antipsychotic medications are recommended for schizophrenia spectrum disorders, yet their long-term effects on functional recovery remain unclear, with conflicting evidence often derived from between-subject comparisons vulnerable to confounding by indication.

**Methods:** We conducted a nationwide register-based cohort study of 65,630 individuals with incident schizophrenia spectrum disorders in Denmark (1998–2023). We modeled antipsychotic exposure against ‘productive engagement’ (employment or education). We used two analytical approaches: (1) within-subject stratified Cox models with time-varying covariates to eliminate time-invariant confounding; and (2) Fine–Gray competing risks models with baseline exposure, accounting for mortality and emigration.

**Results:** Over 26.9 million person-weeks, the overall productive engagement rate was 48.2%. Integration of hospital pharmacy data revealed 6.1% exposure misclassification in studies relying solely on community records. The primary within-subject analysis revealed significant temporal heterogeneity: medication use was associated with reduced engagement rates in the acute (0–2 years; HR = 0.908) and consolidation phases (2–5 years; HR = 0.946), but reversed to a small positive association in the maintenance phase (5+ years; HR = 1.019). The between-subject Fine–Gray model, which estimates cumulative engagement probabilities, yielded an SHR of (95% CI = 0.988–1.015), a population-level average that obscured these phase-specific dynamics.

**Conclusions:** Antipsychotic pharmacotherapy exerts a time-dependent, biphasic impact on vocational recovery. We identified a window of vulnerability during the post-acute ‘consolidation’ phase (years 2–5) where treatment is associated with a transient reduction in productive engagement, before becoming protective after 5 years. These findings challenge the assumption that symptomatic stability automatically facilitates functional reintegration.

**Danish Lay Summary:** I denne undersøgelse har vi set på, hvordan personer kommer tilbage i arbejde eller uddannelse, efter de er startet i medicinsk behandling for skizofreni. Formålet er at forstå, om antipsykotisk medicin øger eller mindsker deres mulighed for at vende tilbage til arbejde eller uddannelse. Resultaterne peger på et udviklingsforløb, som kan forklares i tre faser:

- De første 2 år har en person i medicinsk behandling 9 % lavere sandsynlighed for at være i arbejde eller uddannelse, sammenlignet med perioder hvor personen er uden behandling.
- Efter 2–5 år bliver forskellen mindre (omkring 5 %). Medicinen forsinker stadig tilbagevenden til arbejdsmarkedet eller uddannelse, men i mindre grad end før.
- Efter 5–10 år vender billedet. Her ses en tendens til, at flere som får medicinsk behandling er i arbejde eller uddannelse. Effekten er lille, men den går i en positiv retning.

På kort sigt kan antipsykotisk medicin begrænse muligheden for arbejde eller uddannelse. På længere sigt kan medicinen give den stabilitet, der gør det lettere at fastholde eller vende tilbage til arbejde eller uddannelse.

## 1. Introduction

Schizophrenia spectrum disorders (SSDs) profoundly disrupt employment and social functioning. Despite antipsychotic medications being effective for symptom control, only 32.5% of individuals with first-episode psychosis maintain competitive employment (i.e., open-market paid work or formal education) long-term (Ajnakina et al., 2021), though rates vary considerably across countries and healthcare systems, from below 10% to over 30% depending on welfare context and labor market structure (Marwaha and Johnson, 2004; Holm et al., 2021). Current guidelines recommend continuous maintenance antipsychotic therapy based on strong evidence that discontinuation increases relapse risk (Keepers et al., 2020; Leucht et al., 2012). However, this recommendation prioritizes symptomatic stability over functional outcomes, and clinicians must weigh relapse prevention against potential adverse effects that could impair work capacity (Cornblatt et al., 2011). Recent trials suggest dose reduction strategies may yield comparable or superior long-term functional outcomes despite higher initial relapse rates (Wunderink et al., 2013; Moncrieff et al., 2023).

This study builds on Stürup et al. (2023), who examined antipsychotic patterns and employment outcomes in first-episode schizophrenia using discrete exposure windows (years 2-5) and outcomes at year 6. Stürup et al. (2023) found that individuals who discontinued antipsychotics had higher odds of employment compared with continuous users. We extend this work using a rigorous methodological triangulation approach. We contrast a within-subject design (which controls for all time-invariant genetics and history) against a standard between-subject competing risks analysis. This dual approach enables us to test whether findings from standard observational models hold up when strictly controlling for unmeasured confounding factors, such as illness severity and premorbid cognitive capacity, which are typically prevalent in psychiatric epidemiology.

## 2. Methods

### Study Design and Setting

A protocol with statistical analysis plan was uploaded to medRxiv on October 2, 2025 (Twumasi et al., 2025). Exploratory analysis and preprocessing were carried out before protocol posting. All inferential statistics and survival analysis were completed after the preregistration was accepted on October 3, 2025. Our only departure from the protocol was the decision to include all individuals rather than restrict to 18+. This was decided because the result of productive engagement encompasses both employment and education, making the age restriction to 18+ unnecessary and potentially biasing against cases with onset in adolescents.

We conducted a nationwide, register-based cohort study using a within-subject design in Denmark. Denmark’s tax-funded welfare system ensures universal healthcare access and broad social support, with active labor market policies providing subsidized vocational training (Pedersen et al., 2025) and workplace accommodations for individuals with mental health conditions. The Danish Civil Registration System assigns a unique personal identification number (CPR number) to every resident, enabling unambiguous individual-level linkage across all national registers with complete follow-up and no possibility of opting out (Nordfalk and Hoeyer, 2017), thereby limiting selection bias.

### Data Sources

Data were drawn from multiple validated national registers. The Danish National Patient Register (Plana-Ripoll et al., 2025; Lynge et al., 2011) was used to identify the study cohort and all psychiatric diagnoses. Antipsychotic medication use was ascertained from the Danish National Prescription Register (NPR)(Pottegård et al., 2017), which contains detailed records of all prescriptions dispensed from community pharmacies since 1995. For the period from 2018 onwards, NPR data were supplemented with data from the Danish National Hospital Medicine Register (Sygehusmedicinregisteret, SMR)(Andersen et al., 2024), which records medications administered during hospital stays. Information on labor market status was obtained from the Danish Register for Evaluation and Marginalization (DREAM)(Hjollund et al., 2007), which contains weekly data on all public transfer payments (e.g., sickness benefits, unemployment benefits, disability pension). The DREAM register’s unique weekly granularity in capturing benefit receipt, combined with Denmark’s comprehensive national health registers and universal Central Person Register (CPR) number linkage system, creates a unique opportunity to examine the longitudinal association between medication patterns and labor market outcomes in schizophrenia spectrum disorders.

### Data availability and guidelines

This study used Danish register data provided by Statistics Denmark. Access to data can be granted for approved research projects. All microdata were handled according to guidelines to prevent identification, including rounding counts. No additional data are publicly available.

### Study Population

The source population included all individuals residing in Denmark who received a first-time diagnosis of a nonaffective psychotic disorder (ICD-10 codes F20-F29). The date of the first registered F20-F29 diagnosis was defined as the cohort entry date.

To ensure complete temporal alignment between medication exposure and employment data, we restricted the cohort to individuals with index diagnosis dates between January 1, 1998, and December 31, 2023, reflecting the period for which relevant DREAM labor market affiliation data were available for this project.

Individuals were excluded if they died or emigrated within 2 years of cohort entry. This 2-year exclusion filter removed 1,000 patients (1.52% of the cohort). Community pharmacy records (NPR) do not capture medications administered during inpatient stays, or free medication offered to patients with recent onset schizophrenia (Jensen et al., 2020), which could lead to exposure misclassification bias in our main sample. Therefore, a further subgroup analysis on SMR data, which includes the first 2 years of prescriptions for members of the cohort first diagnosed after 2018, was completed.

### Study Variables Antipsychotic Exposure

The primary exposure was antipsychotic medication use, modeled as a time-varying variable. For the period 1998–2017, exposure was based on filled prescriptions in the NPR (ATC code N05A, excluding lithium N05AN01). For the 2018–2023 sensitivity analysis, exposure was based on an integrated record of NPR prescriptions and SMR in-hospital prescriptions Andersen et al. (2024).

We calculated medication exposure duration for each prescription using Tiihonen’s methodology Tiihonen et al. (2011), adapted from the original protocol used in the nationwide Finnish schizophrenia cohort study. This approach models individual prescription coverage periods based on package characteristics and Defined Daily Doses (DDDs) from the World Health Organization ATC/DDD Index 2025 (WHO, 2025).

For each prescription, exposure duration in days was calculated as:

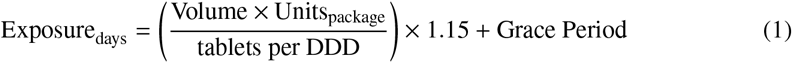

where Volume denotes the number of packages dispensed, Units_package_ the discrete count of dosage units (e.g., tablets) per package as recorded in the prescription register variable *strnum*, and tablets per DDD the number of units required to achieve one Defined Daily Dose as specified by the WHO ATC/DDD Index. The 1.15 multiplier accounts for dosing variability and patient stockpiling behavior Tiihonen et al. (2011). Grace periods differed by formulation: 14 days for oral medications and 28 days for long-acting injectable (LAI) depot formulations, reflecting typical administration intervals. Calculated durations were capped at 7-180 days per prescription to exclude implausible values.

Person-weeks were classified as *AP use* if they fell within an active prescription coverage period, and *no AP use* otherwise. This duration-based approach provides more accurate exposure estimation than simple unit-counting methods, particularly for LAI formulations where package size does not reflect duration of action.

Antipsychotics were further categorized by generation: *first-generation* (typical) antipsychotics included ATC codes N05AA, N05AB, N05AC, N05AD, N05AF, and N05AG; *second-generation* (atypical) antipsychotics included N05AE, N05AH, N05AL, and N05AX. Clozapine (N05AH02) was analyzed separately given its unique indication for treatment-resistant schizophrenia. LAI formulations were identified through product name keyword matching.

When multiple medications were dispensed concurrently within a person-week, classification prioritized clozapine, followed by LAI, then oral antipsychotics. This hierarchy reflects clinical severity: clozapine is reserved for treatment-resistant disease, LAIs are typically prescribed for patients with adherence difficulties, and oral antipsychotics represent first-line treatment. Polypharmacy was defined as concurrent use of both a first-generation and a second-generation antipsychotic within the same person-week, representing cross-class combination therapy that may indicate greater treatment complexity or illness severity. Medication switching was defined as any transition between exposed and unexposed states during follow-up, with individuals contributing person-time to both states classified as switchers.

### Hospital Medication Data Integration

For the subcohort diagnosed from 2018 onwards (N=14,190, 30% of total cohort), we integrated hospital pharmacy records from the SMR Andersen et al. (2024) to eliminate exposure misclassification bias in the early post-diagnosis period. SMR data contained 1,218,821 antipsychotic prescriptions (ATC code N05A minus lithium). The SMR contained true dose strength data (99.99% complete) enabling more accurate DDD calculations using the following

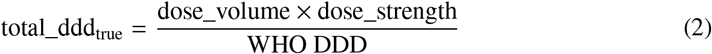

where grace period = 14 days (oral) or 28 days (LAI).

### Outcome Measures

The primary outcome was ‘productive engagement’, a time-varying binary variable at the person-week level indicating whether an individual was engaged in competitive employment, formal education, or a vocational training program. The operationalization was based on a hierarchical classification of weekly labor market affiliation, with specific codes from the DREAM used to define each state (see Supplementary Table 1 Appendix A for full details).

### Covariates

The within-subject design controlled for all time-invariant confounders. The analysis adjusted for two key time-varying covariates that were updated at each person-week observation: age and recent hospital discharge. Finally, a baseline time-invariant covariate, gender (legal sex represented by CPR number), was also included in all models. Time-varying covariate adjustment was implemented using the counting process formulation of Cox proportional hazards regression models (in the following termed ‘Cox models’) within a person-week panel data structure. Each individual contributed multiple observations (one per week of follow-up), with covariate values updated at each time point. The models used within-person stratification which conditions on the individual-level baseline hazards and effectively controls for all time-invariant unmeasured confounders (equivalent to a fixed-effects approach).

### Statistical Analysis

We employed methodological triangulation (Lawlor et al., 2017) using multiple analytical approaches with different assumptions to allow us to attempt to understand potential associations beyond what a single method could offer. This strategy addresses the fundamental challenge that all observational pharmacoepidemiologic analyses are vulnerable to unmeasured confounding, with different methods susceptible to different biases. By examining whether associations persist across methods with non-overlapping assumptions. Our triangulation strategy combined: (1) within-subject stratified Cox models controlling for time-invariant confounding through self-comparison, (2) time-stratified analyses characterizing temporal heterogeneity in medication effects, (3) lagged exposure analyses addressing reverse causation and delayed effects, and (4) Fine–Gray competing risks models (Fine and Gray, 1999) accounting for mortality and emigration as competing events.

### Primary Analysis: Within-Subject Stratified Cox Models

The primary analysis was restricted to 50,440 individuals who contributed person-time to at least two different antipsychotic exposure categories (i.e., “switchers”), enabling within-subject comparisons. We used stratified Cox models to estimate hazard ratios (HRs) and 95% confidence intervals (CIs) for the association between changes in antipsychotic exposure status and productive engagement transitions. In this model, each individual serves as their own control, comparing rates of productive engagement during medication-exposed versus unexposed weeks within the same person. This design eliminates confounding from all time-invariant factors including genetics, childhood adversity, baseline cognitive ability, and premorbid functioning.

This within-subject design requires patients to have variation in medication status during follow-up; 15,190 individuals (23.14%) with invariant medication patterns were excluded from Cox models but retained for descriptive analyses. Nonswitchers could represent clinically distinct subgroups (severe treatment-adherent patients versus treatment-refusing individuals), with exclusion potentially limiting generalizability to patients with stable medication patterns.

### Time-Stratified Analysis

The proportional hazards assumption underlying Cox models requires that hazard ratios remain constant over follow-up time. We assessed this assumption using Schoenfeld (1982) residuals tests for all models. In register-based studies of this magnitude, Schoenfeld tests typically reject the null hypothesis of proportional hazards even with minor deviations, due to the substantial statistical power afforded by large sample sizes. We therefore treated statistically significant violations as an indication of potentially meaningful temporal heterogeneity warranting further investigation through time-stratified analyses, rather than as evidence that Cox models are fundamentally unsuitable.

We computed time-stratified Cox models as the analytical approach when PH violations were detected. The 10-year follow-up was divided into three clinically meaningful periods from first diagnosis date (index date): 0–2 years (acute/early recovery phase), 2–5 years (consolidation phase), and 5+ years (long-term maintenance phase). Separate Cox models were fitted within each stratum, allowing hazard ratios to vary across periods while maintaining within-person control for time-invariant confounding. This approach provides period-specific effect estimates characterizing how medication-employment associations evolve across the recovery trajectory.

### Secondary Analysis: Fine–Gray Competing Risks (Between-Subject)

To provide a prognostic benchmark comparable to standard observational literature, we employed Fine–Gray subdistribution hazard models (Fine and Gray, 1999; Austin and Fine, 2017). Critically, unlike the primary within-subject Cox models, the Fine–Gray analysis was a between-subject design using baseline exposure (medication status at first diagnosis) and baseline covariates (age, gender), treating death and emigration as competing events that preclude productive engagement. This model estimates the cumulative probability of employment for the full cohort (N=65,630) but does not control for unmeasured time-invariant confounding or treatment switching, these are vulnerabilities that the within-subject Cox design eliminates. We interpreted discrepancies between these models through the lens of causal hierarchy: the within-subject Cox model was prioritized for etiological inference due to its superior control for unmeasured confounding, while the Fine–Gray model served as a ‘real-world’ prognostic reference reflecting the combined impact of treatment assignment, adherence patterns, and selection bias inherent in standard population comparisons. Importantly, subdistribution hazard ratios from the Fine–Gray model do not possess a simple rate-ratio interpretation analogous to cause-specific hazard ratios (Andersen et al., 2012); they reflect the relative ordering of cumulative incidence curves rather than a traditional epidemiological rate.

### Sensitivity Analyses

For the 2018+ subcohort with hospital pharmacy data availability (N=14,190), we compared employment outcomes under three medication exposure definitions: (1) community pharmacy only, (2) hospital pharmacy only, and (3) combined community and hospital coverage. This quantifies exposure misclassification bias magnitude from incomplete medication ascertainment during hospitalization periods when community pharmacy records underestimate treatment intensity.

We examined effect heterogeneity across 8 dimensions (27 strata total): gender, age at diagnosis, geographic region, diagnosis subtype, medication adherence, hospitalization type (inpatient/outpatient/emergency), and SMR data availability (pre-2018 vs 2018+).

### Model Specifications

Six Cox proportional hazards models were fitted to examine different aspects of antipsychotic medication effects on productive engagement. Models 1 and 2 constituted the primary analyses; Models 3–6 were prespecified exploratory analyses. Model 1 examined any antipsychotic use versus no use, adjusting for age, recent hospital discharge, and gender. Model 2 assessed antipsychotic polypharmacy versus monotherapy or no treatment, with the same covariate adjustment. Model 3 evaluated a four-level medication type classification (No AP, Clozapine, LAI, Oral AP) incorporating time-varying covariates for age and recent discharge status. Model 4 tested a dose–response relationship using total exposure days (0–7 days per week) restricted to person-weeks with antipsychotic use. Model 5 attempted to fit a time-varying coefficient model (Therneau et al., 2017) allowing the effect of antipsychotic use to vary with log-transformed follow-up time; this model failed to converge due to the scale of the dataset. We therefore fitted an interaction model testing whether medication effects differed during the acute post-discharge period versus stable community residence.

To examine delayed medication effects and address potential reverse causation, we conducted lagged exposure analyses with 6-month, 12-month, and 24-month delays between exposure measurement and outcome assessment. By allowing time between exposure and outcome windows, these analyses reduce the risk that employment status influences medication adherence rather than vice versa, while testing the hypothesis that medication effects on employment emerge gradually over extended treatment periods.

The secondary Fine–Gray analysis (described above) estimated subdistribution hazard ratios (SHR) for comparison with the primary within-subject Cox models.

To characterize treatment complexity, we mapped all medication switches using Sankey flow diagrams, identifying transitions between medication types (first-generation only, second-generation only, clozapine, polypharmacy, and no AP use).

### Multiple Testing Correction

Benjamini-Hochberg (Benjamini and Hochberg, 1995) False Discovery Rate correction at the 5% threshold was applied to exploratory subgroup and sensitivity analyses examining effect heterogeneity across clinical and demographic strata.

### Software

Data preparation was carried out in SAS (version 9.4), and Stata (version 18). Analysis was carried out using R version 4.5.1, with packages survival (Therneau, 2024), survminer (Kassambara et al., 2021), on secure Statistics Denmark servers.

### Ethics

The study was approved by the Regional Data Protection Agency (P-2020-88). The authors assert that all procedures contributing to this work comply with the ethical standards of the relevant national and institutional committees on human experimentation and with the Helsinki Declaration of 1975, as revised in 2008.

## 3. Results

### Study Cohort and Follow-up

The final analytic cohort comprised 65,630 individuals with incident schizophrenia spectrum disorders, contributing 26.9 million person-weeks of observation over a maximum analytical follow-up of 10 years (median 10.00 years, IQR: 6–10 years). By design, all estimates are representative of individuals who survived and did not emigrate during the first 2 years following diagnosis. Baseline characteristics are described in Table 1. The cohort was predominantly male (51.2%), with median age 30 years at first diagnosis. Notably, 82.9% reached the maximum 10-year follow-up, 81.6% received at least one antipsychotic prescription during follow-up, and 76.9% (n=50,440) were classified as medication switchers eligible for within-subject Cox models.

**Table 1:** Baseline Characteristics of Study Cohort.

| Characteristic | Overall Cohort<br>(N=65,630) |
| --- | --- |
| **DEMOGRAPHICS** |  |
| Age at diagnosis, median (IQR), years | 30 (21–47) |
| Age at diagnosis, mean (SD), years | 36.0 (18.9) |
| 10–15 years, n (%) | 6,940 (10.6) |
| 15–20 years, n (%) | 18,010 (27.5) |
| 20–25 years, n (%) | 13,880 (21.2) |
| 25–30 years, n (%) | 9,170 (14.0) |
| 30–35 years, n (%) | 7,020 (10.7) |
| 35+ years, n (%) | 10,610 (16.2) |
| Male gender, n (%) | 33,620 (51.2) |
| Female gender, n (%) | 32,000 (48.8) |
| **PRIMARY DIAGNOSIS (ICD-10 F20-F29)** |  |
| F23 Acute transient psychotic disorders, n (%) | 16,950 (25.8) |
| F20 Schizophrenia, n (%) | 16,370 (24.9) |
| F22 Persistent delusional disorders, n (%) | 11,120 (16.9) |
| F21 Schizotypal disorder, n (%) | 9,280 (14.1) |
| F29 Unspecified nonorganic psychosis, n (%) | 8,440 (12.9) |
| F28 Other nonorganic psychotic disorders, n (%) | 1,830 (2.8) |
| F25 Schizoaffective disorders, n (%) | 1,610 (2.5) |
| F24 Induced delusional disorder, n (%) | 40 (0.1) |
| **FOLLOW-UP** |  |
| Median follow-up, years (IQR) | 10 (6–10) |
| Mean follow-up, years (SD) | 7.85 (2.9) |
| **CENSORING EVENTS** |  |
| Reached 10-year cap, n (%) | 54,410 (82.9) |
| Death, n (%) | 9,690 (14.8) |
| Emigration, n (%) | 1,540 (2.3) |
| **MEDICATION EXPOSURE (DURING FOLLOW-UP)** |  |
| Any antipsychotic use, n (%) | 53,530 (81.6) |
| Never used antipsychotics, n (%) | 12,110 (18.4) |
| Person-week coverage rate, % | 40.7 |
| Polypharmacy (≥2 AP classes), % of weeks | 1.4 |
| **PRODUCTIVE ENGAGEMENT** |  |
| Overall rate during follow-up, % | 48.2 |
| **BASELINE EMPLOYMENT STATUS** |  |
| Employment, n (%) | 21,930 (33.4) |
| Unemployment, n (%) | 7,660 (11.7) |
| Education/training, n (%) | 4,230 (6.5) |
| Social assistance, n (%) | 2,010 (3.1) |
| Sickness absence, n (%) | 1,610 (2.5) |
| Retirement/pension, n (%) | 960 (1.5) |

Table 1 presents baseline employment status at diagnosis (33.4% employed), while the 48.2% productive engagement rate represents the longitudinal proportion across all person-weeks during follow-up, reflecting cumulative engagement over time rather than point prevalence at cohort entry.

Censoring flow patterns are visualized in Supplementary Figure A.7.

### 3.1. Primary Analysis (Within-Subject Cox)

Time-stratified within-subject Cox models revealed distinct phase-specific effects on productive engagement (Figure 3). Schoenfeld residual tests indicated proportional hazards violations across all models (all p<0.001), confirming temporal heterogeneity that warranted period-specific estimation.

In the acute phase (0–2 years, 3.1 million person-week events), antipsychotic use was associated with a 9% reduction in the rate of productive engagement (HR=0.908, 95% CI: 0.903– 0.913). This negative association persisted but attenuated during the consolidation phase (2–5 years, 3.5 million events: HR=0.946, 95% CI: 0.939–0.952). Crucially, in the long-term maintenance phase (5+ years, 3.2 million events), the direction of effect reversed, with medication showing a modest positive association with employment (HR=1.019, 95% CI: 1.010–1.028).

These within-subject estimates compare medication-exposed versus unexposed person-weeks within the same individual, thereby controlling for all time-invariant confounders including genetics, premorbid functioning, and baseline illness severity. The J-shaped pattern: early negative associations reversing to late positive associations, represents the principal within-subject finding of this study.

### 3.2. Secondary Analysis (Fine–Gray Competing Risks)

The between-subject Fine–Gray analysis, using baseline medication exposure (at first diagnosis) and adjusting only for age and gender, yielded a null result (SHR=1.002, 95% CI: 0.988–1.015). This suggests that, when viewing the population as static groups defined at diagnosis, the ‘treated’ and ‘untreated’ cohorts achieved similar cumulative productive engagement probabilities (Andersen et al., 2012).

However, given the minimal covariate adjustment compared to the within-subject Cox models, this null finding likely reflects the cancellation of opposing forces: the drug’s stage-specific pharmacological effects masked by confounding by indication, where more severely ill patients are selectively prescribed antipsychotics. Over a 10-year follow-up, 38,160 individuals (69.1%) achieved productive engagement at least once, 11,730 (21.2%) remained alive but never engaged, and 550 (1.0%) experienced competing events (death or emigration).

The divergence between the within-subject Cox findings (which revealed phase-specific HRs ranging from 0.908 to 1.019) and the Fine–Gray result (SHR=1.002) should be interpreted cautiously, as these models estimate fundamentally different quantities (Andersen et al., 2012): the Cox model yields cause-specific rate ratios among those at risk, whereas the subdistribution hazard ratio reflects the ordering of cumulative incidence curves and lacks a simple rate-ratio interpretation. Nonetheless, the contrast underscores that between-subject comparisons using baseline exposure cannot resolve the stage-specific dynamics that emerge from within-subject longitudinal designs.

### Exploratory Subgroup Analyses

The following exploratory analyses further characterize temporal patterns; these patterns should be interpreted as hypothesis-generating.

Cox model results are presented across four forest plots: Figure 1 (medication types by period), Figure 2 (dose–response), Figure 3 (time-stratified overall effects), and Figure 4 (lagged exposure). Each forest plot displays hazard ratios with 95% confidence intervals, with the vertical dashed line at HR=1.0 indicating no effect.

**Figure 1:**
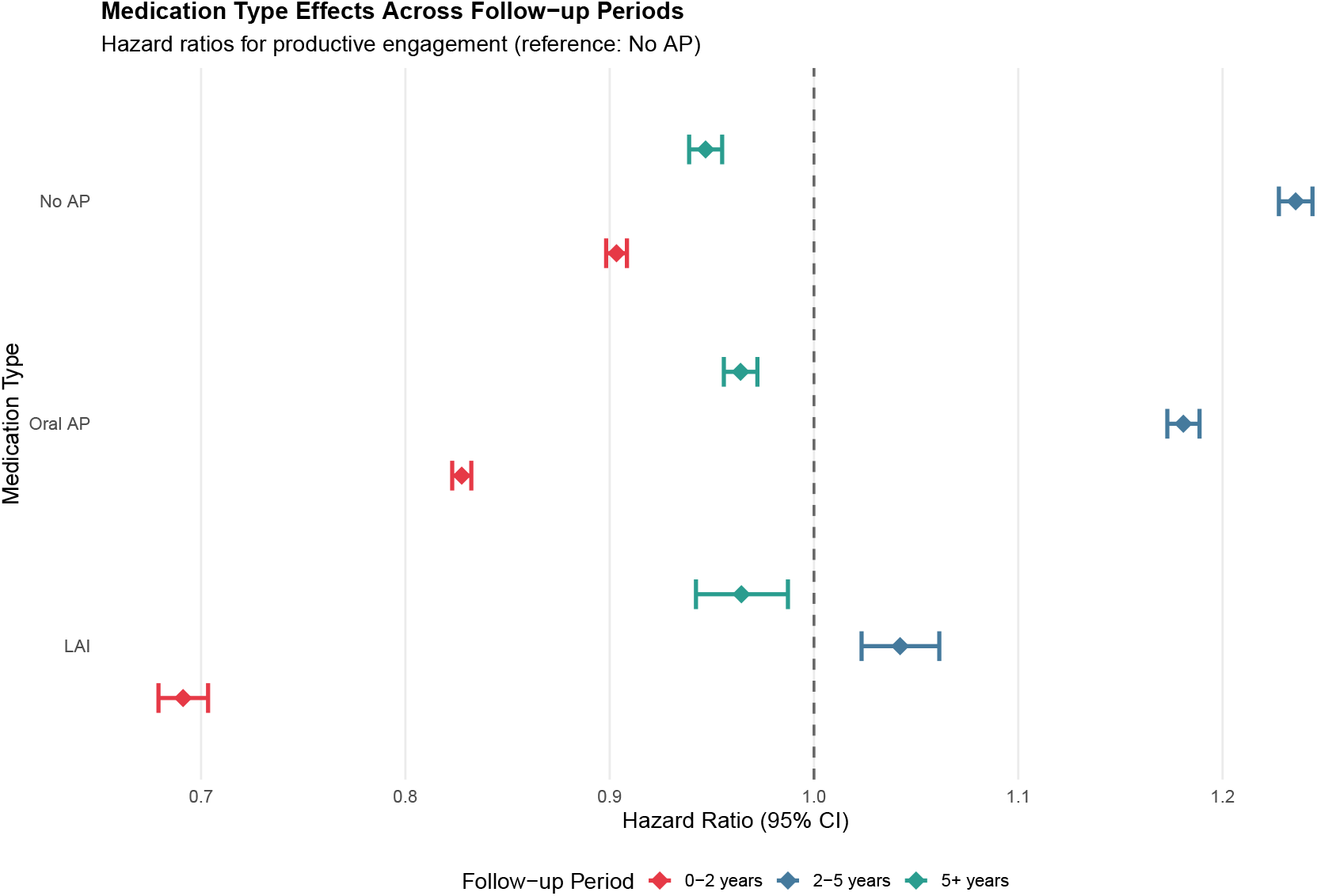
Medication type effects vary dramatically across follow-up periods (within-subject analysis). Hazard ratios from within-person stratified Cox models comparing medication-exposed versus unexposed person-weeks within the same individual.

**Figure 2:**
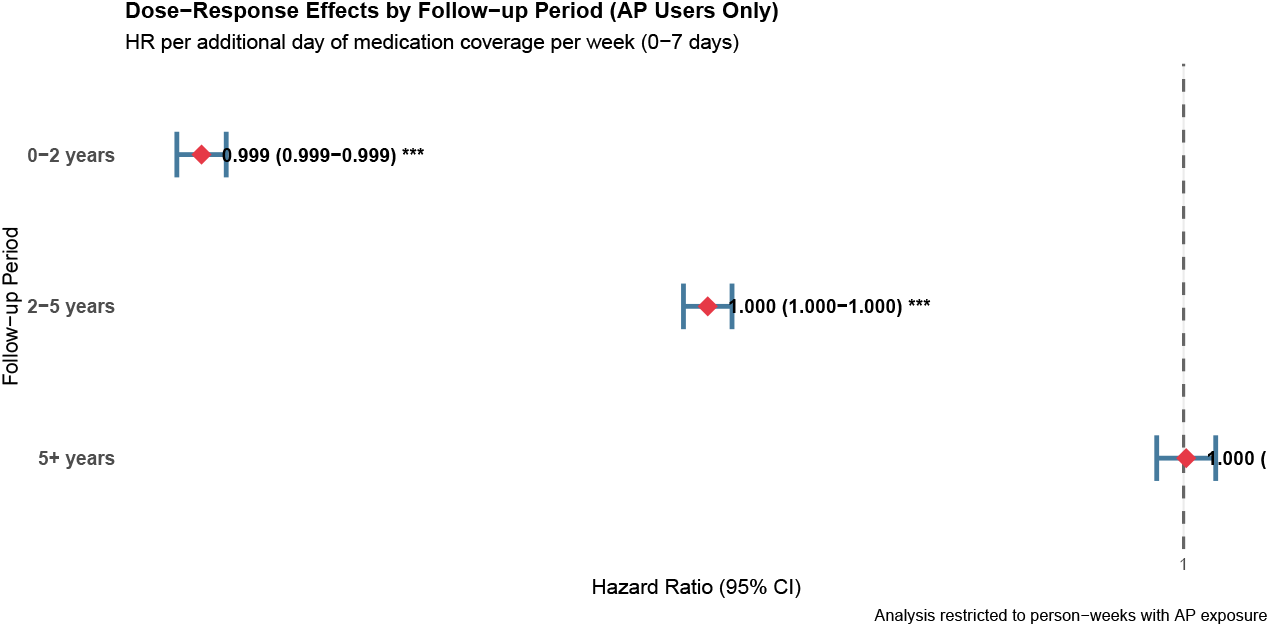
Dose–response analysis reveals minimal incremental effects of medication coverage intensity.

**Figure 3:**
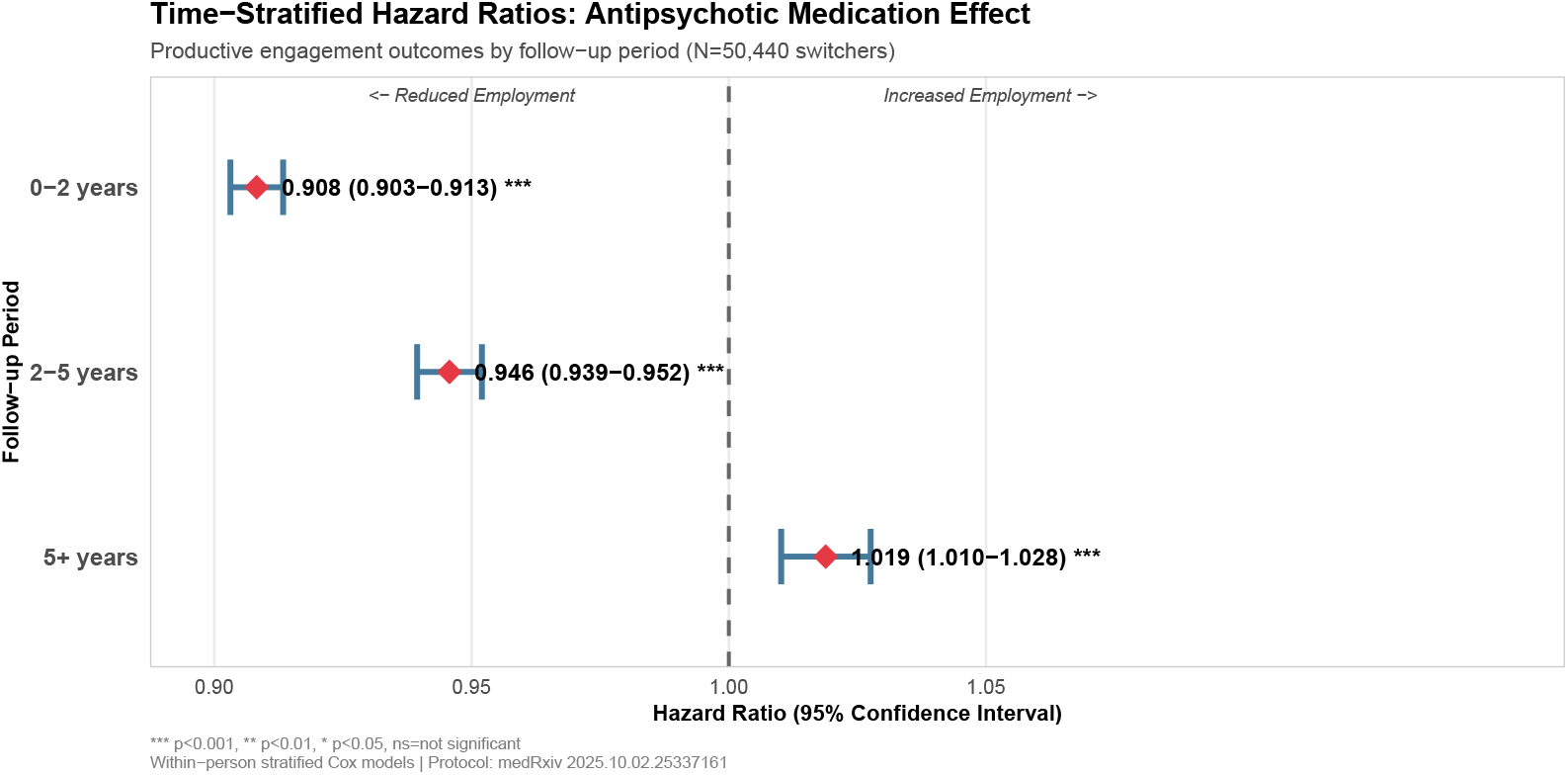
Time-stratified hazard ratios reveal temporal heterogeneity (within-subject stratified analysis). Each patient serves as their own control, eliminating time-invariant confounding.

**Figure 4:**
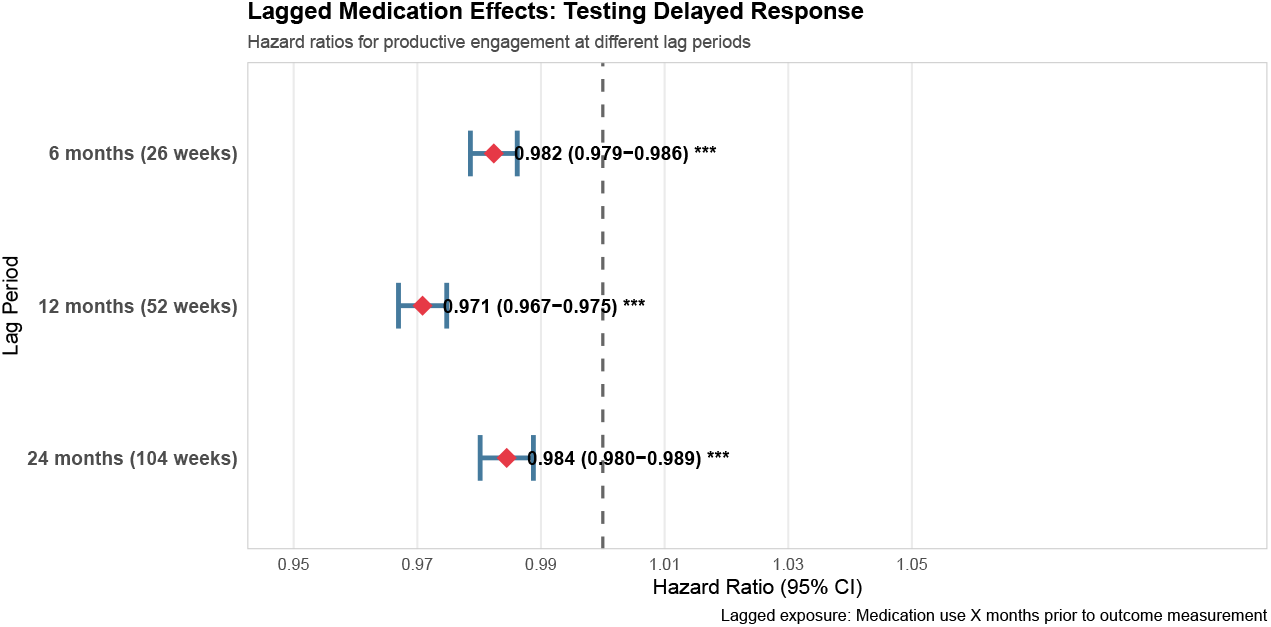
Lagged exposure analysis reveals persistent negative associations (within-subject analysis). Negative associations persist across 6-, 12-, and 24-month lag periods, suggesting findings are not fully explained by reverse causation.

Medication type effects exhibited temporal heterogeneity (Figure 1). During the acute phase (0–2 years), long-acting injectables showed the strongest negative associations (LAI: HR=0.691, 95% CI: 0.679-0.705), while clozapine (HR=0.903, 95% CI: 0.880-0.927) and oral antipsychotics (HR=0.828, 95% CI: 0.823-0.832) showed intermediate effects. All three medication types reversed to positive associations during consolidation (2–5 years), with clozapine showing the strongest positive association (HR=1.236, 95% CI: 1.210-1.261). The late phase (5+ years) showed a second reversal to modest negative associations across all formulations (HRs 0.947-0.964).

Dose–response analyses among person-weeks with antipsychotic exposure revealed essentially null findings across all temporal periods (Figure 2). Hazard ratios per additional day of weekly coverage ranged from 0.989 to 1.000 across periods (95% CIs extremely tight due to sample sizes exceeding 2-4 million person-weeks per period), with no clinically meaningful dose–response gradient. The absence of dose–response effects suggests that medication presence matters more than coverage intensity for employment outcomes, consistent with threshold effects rather than linear dose–response relationships. Note that these dose–response analyses were restricted to person-weeks with antipsychotic exposure, examining intensity effects within treated periods; this does not contradict differential effects between medication categories or between exposed versus unexposed states observed in time-stratified models.

### Medication Exposure Patterns

Using Tiihonen methodology, antipsychotic coverage was observed in 40.65% of all person-weeks, with second-generation agents predominating (32.41%).

### Sensitivity Analysis 1: Lagged Exposure Effects

Lagged exposure analyses testing temporal delays between medication exposure and employment outcomes revealed consistent negative associations across all delay periods, though with temporal attenuation suggesting medication-employment relationships evolve with treatment duration (Figure 4). At 6-month lag (13.9 million person-weeks), medication exposure 6 months prior associated with modest reduced engagement (HR=0.982, 95% CI: 0.979-0.986). The 12-month lag demonstrated the weakest association (HR=0.971, 95% CI: 0.967-0.975). At 24-month lag, the association remained negative but attenuated to the 6-month level (HR=0.984, 95% CI: 0.980-0.989, 16.0 million person-weeks).

The weakest association at 12-month lag aligns with time-stratified findings that medication-employment associations attenuate gradually over the 2–5 year consolidation period. The persistence of negative associations across all lag periods argues against reverse causation (unemployment triggering medication initiation) and is consistent with delayed medication effects that attenuate gradually as functional recovery trajectories unfold.

### 3.3. SMR Hospital Medication Integration: Quantifying Exposure Misclassification Bias

For the 2018+ subcohort with hospital pharmacy data availability (N=14,190), integration of Sygehusmedicinregisteret (SMR) records with community prescriptions substantially reduced exposure misclassification bias. Community pharmacy coverage alone captured 26.69% of person-weeks (1,173,836 weeks), while SMR hospital pharmacy captured an additional 14.94% (657,012 weeks). Combined community and SMR coverage reached 33.25% (1,465,012 weeks), yielding an absolute coverage gain of 6.56% (291,176 person-weeks) and relative increase of 24.6%. Notably, 291,176 weeks (44.3% of SMR coverage, 6.56% of all 2018+ person-weeks) had SMR-only coverage with no concurrent community dispensing, representing medication exposure periods that would have been misclassified as unexposed under community pharmacy data alone, particularly during early post-diagnosis hospitalization periods.

Cox models comparing exposure definitions quantified the resulting bias. Community-only methodology yielded HR=0.96 (95% CI: 0.95-0.96), while combined community and SMR methodology strengthened the association to HR=0.90 (95% CI: 0.89-0.91), demonstrating 6.09% exposure misclassification bias (HR ratio: 0.96/0.90=1.067). Relying solely on community pharmacy records systematically underestimates medication-employment associations by failing to capture hospital-administered medications during acute treatment episodes. SMR integration eliminates this bias for the 2018+ cohort, providing complete medication ascertainment across hospital and community settings (Figure 5).

**Figure 5:**
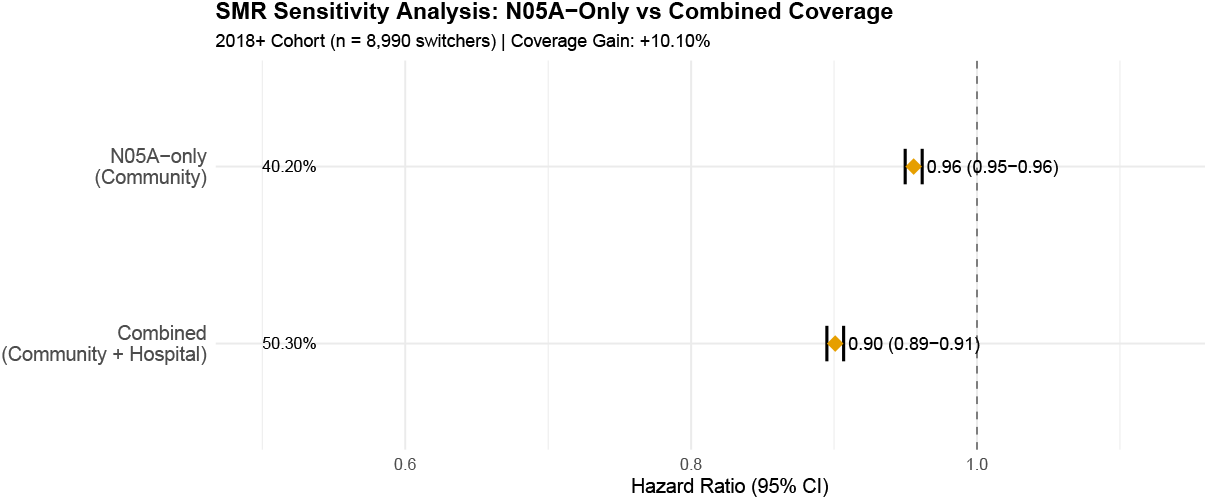
SMR hospital medication integration reveals 6.09% exposure misclassification bias.

This bias is particularly relevant for the early post-diagnosis period when hospitalization rates are highest. For pharmacoepidemiologic studies of psychotic disorders, these findings demonstrate that community pharmacy data alone systematically underestimate medication exposure, biasing effect estimates toward the null.

### Overall Cox Models (Supplementary Materials)

Overall pooled Cox models (HR=1.08 for AP use, HR=0.70 for polypharmacy) are provided in A.8 but should be interpreted with caution. Severe proportional hazards violations (p<0.0001) indicate these estimates mask temporal heterogeneity, averaging across periods with divergent effects (0–2 yr: HR=0.908, 2–5 yr: HR=0.946, 5+yr: HR=1.019). Given these violations, the polypharmacy estimate (HR=0.70) should not be interpreted as a stable treatment effect; time-stratified analyses provide more reliable period-specific estimates.

### Medication Switching Patterns

Among the 51,720 individuals who made medication category transitions during follow-up, treatment trajectories revealed substantial heterogeneity (Figure 6). The majority of transitions occurred between second-generation antipsychotics (68% of all transitions). Discontinuation events accounted for 22% of all transitions, with patients moving from medicated to unmedicated states. Notably, 8.2% of the cohort received clozapine during follow-up, representing treatment change for individuals with treatment-resistant schizophrenia.

**Figure 6:**
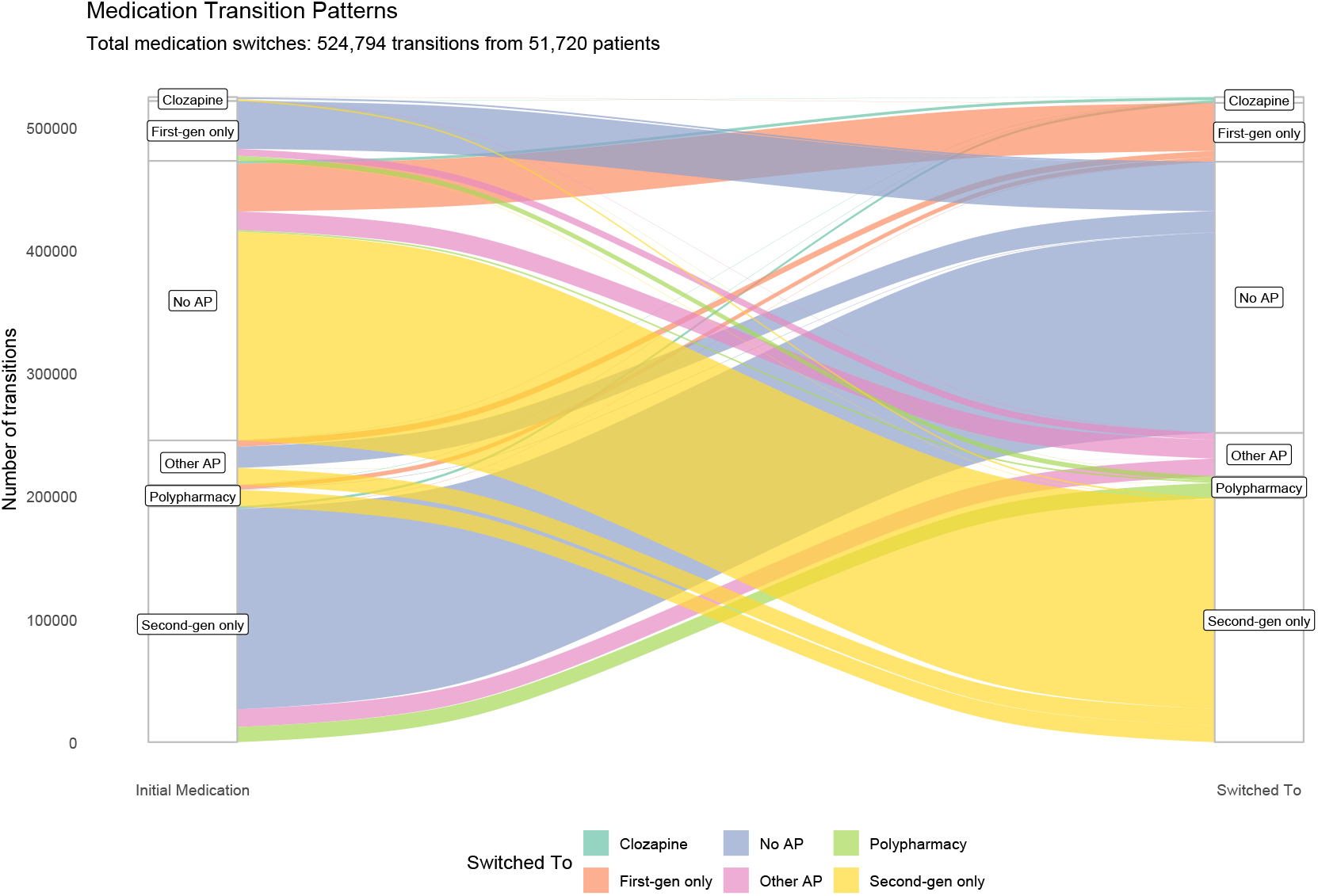
Sankey diagram showing medication switching patterns among switchers (n=50,440). Flow width proportional to number of transitions. Medication categories: first-generation only, second-generation only, clozapine, polypharmacy, and no AP use.

## 4. Discussion

### 4.1. Principal Findings

In this nationwide cohort of 65,630 individuals with schizophrenia spectrum disorders, we demonstrate that the impact of antipsychotic medication on vocational recovery is not static but highly stage-dependent. Using a within-subject design that controlled for genetic and historic confounding, we found that medication was associated with reduced productive engagement during the acute phase (0–2 years: HR=0.908, 95% CI: 0.903–0.913) and the consolidation phase (2–5 years: HR=0.946, 95% CI: 0.939–0.952), before reversing to a small positive association in the long-term maintenance phase (5+ years: HR=1.019, 95% CI: 1.010–1.028). While the late-phase HR of 1.019 is extremely small in magnitude, its clinical relevance could be offered in two features: first, applied across millions of person-weeks in a population-level analysis, even small effect sizes translate to meaningful differences in cumulative employment outcomes; second, the pattern, which is a reversal from negative to positive association is more informative than a small single point estimate, suggesting a potential shift in the medication-employment relationship over the recovery trajectory. The between-subject Fine–Gray competing risks analysis yielded an SHR of 1.002, suggesting equivalent cumulative engagement probabilities for baseline-treated and untreated groups. However, the subdistribution hazard ratio does not possess a simple rate-ratio interpretation analogous to the cause-specific hazard ratios from our Cox models (Andersen et al., 2012), and these two quantities should not be directly compared. The discrepancy between models reflects three overlapping factors: first, the Fine–Gray SHR and the cause-specific HR estimate fundamentally different statistical quantities; second, the between-subject design is vulnerable to confounding by indication, where patients with more severe baseline illness are selectively prescribed antipsychotics; and third, the baseline exposure definition cannot capture treatment switching over follow-up. Together, these factors highlight the necessity of within-subject designs for etiological inference regarding medication–employment associations.

### 4.2. The Hierarchy of Evidence: Why Within-Subject Models Matter

The divergence between our primary (within-subject) and secondary (between-subject) analyses offers an important methodological lesson. The Fine–Gray model yielded an SHR near unity, which might superficially suggest no treatment–outcome association. However, as Andersen et al. (2012) demonstrate, the subdistribution hazard ratio reflects the ordering of cumulative incidence curves but its numerical value does not possess a simple epidemiological interpretation. More fundamentally, the between-subject design is vulnerable to confounding by indication: patients with more severe illness are selectively prescribed antipsychotics, and this selection bias attenuates observed associations.

By contrast, our within-subject Cox model eliminates time-invariant confounding by comparing each patient to themselves during ‘on’ and ‘off’ medication periods. The emergence of phase-specific associations (negative in early years, positive in late years) from this more rigorous design demonstrates that averaged between-subject estimates can obscure clinically meaningful temporal heterogeneity. We conclude that for etiological questions in schizophrenia, where unmeasured confounding is pervasive, within-subject designs should be prioritized over standard cohort comparisons (Hernán, 2010; Wolbers et al., 2009; Lau et al., 2017; Andersen et al., 2012).

### 4.3. Clinical Implications: A Stage-Specific Approach

The biphasic trajectory observed in our primary analysis: an initial inverse relationship between treatment and employment followed by long-term stabilization, challenges the static nature of current treatment guidelines. While current protocols rightly prioritize relapse prevention, our data indicates that for the first 5 years of illness, this symptomatic control comes at a functional cost. The negative association observed during the ‘consolidation’ phase (years 2–5; HR 0.946) suggests that once the acute psychotic crisis has resolved, the iatrogenic burden of treatment, potentially manifesting as sedation, cognitive dulling, or secondary negative symptoms may act as a ‘functional ceiling’ that actively suppresses vocational reintegration.

This finding isolates the consolidation phase as a critical period of vulnerability where pharmacotherapy alone is insufficient. Consequently, clinical practice should consider moving beyond a predominantly pharmacological approach during these intermediate years. Vocational rehabilitation should be indicated not merely as a social adjunct, but as a necessary clinical complement to the medication-associated functional dampening identified in this window. The reversal to a protective association in the maintenance phase (5+ years; HR 1.019) validates the necessity of long-term antipsychotic coverage for chronic stability, but it implies that the path to this stability requires active management of the initial functional trade-off. We propose a stage-matched clinical model: intensive functional scaffolding and dose optimization should be front-loaded into the 2–5 year post-diagnosis window to bridge the gap between symptomatic remission and the long-term protective benefits of maintenance treatment.

The differential findings across medication categories warrant cautious interpretation. The strong negative acute-phase association for LAIs (HR=0.691) likely reflects selection bias: LAIs are preferentially prescribed to patients with adherence difficulties, who may have more severe illness and poorer baseline functioning. The positive consolidation-phase association for clozapine (HR=1.236) may similarly reflect treatment-resistant patients achieving clinical stabilization rather than a direct pharmacological effect on employment. These category-specific patterns should be interpreted as reflecting the clinical context of prescribing rather than as direct comparisons of medication efficacy for employment outcomes.

### 4.4. Comparison with Prior Studies

Our within-subject findings contrast with prior work. Solmi et al. (2022) found antipsychotic treatment associated with 30–50% lower risk of work disability (HR≈0.65) in Swedish patients with first-episode nonaffective psychosis, with long-acting injectables showing strongest protective associations. Similarly, Matsuzaki et al. (2023) reported higher employment during clozapine treatment using a mirror-image design. Holm et al. (2021) documented low baseline employment rates (24% for schizophrenia) in Nordic populations, consistent with our 33.4% at diagnosis. Our more granular time-stratified approach reveals that these averaged effects may obscure important temporal heterogeneity: medication effects vary substantially across the recovery trajectory.

### 4.5. Lagged Exposure Analyses

Building upon this premise, it is necessary to explore the potential clinical mechanisms underlying these delayed functional effects. The observation that negative associations persist even when the measurement of exposure is separated from the outcome by up to 2 years implies that the functional consequences of antipsychotic treatment manifest over a prolonged timeline. From a clinical perspective, this delayed impact may reflect the cumulative and compounding burden of medication side effects, such as persistent sedation, cognitive dulling, or the emergence of secondary negative symptoms. These negative factors can gradually erode a patient’s capacity to maintain competitive employment or educational commitments, acting as a persistent functional ceiling even after acute psychotic symptoms have been successfully managed.

However, a balanced interpretation requires acknowledging that lagged models cannot completely disentangle pharmacological effects from the underlying trajectory of the illness. It remains plausible that patients who consistently require antipsychotic medication over a protracted 2-year period represent a distinct subgroup suffering from a more severe or enduring form of the disorder. Under this interpretation, the persistent negative associations observed in our lagged models might still partially capture the chronic, disabling nature of the schizophrenia spectrum itself, rather than reflecting an exclusively iatrogenic pharmacological cost. Therefore, while the lagged exposure framework robustly challenges the hypothesis that acute contemporaneous relapse is the sole driver of vocational loss, the delayed effects we observe most likely represent a complex interaction between the prolonged pharmacological burden of maintenance treatment and the inherent long-term functional decline associated with persistent psychiatric morbidity.

### 4.6. Strengths and Limitations

Our study’s primary strength is the integration of SMR hospital pharmacy data (correcting 6.1% exposure misclassification) combined with the statistical power of 26.9 million person-weeks and a rigorous within-subject design that controls for all time-invariant confounders.

The major limitation of the Fine–Gray analysis was its inability to accommodate time-varying covariates or within-subject stratification, rendering it a descriptive rather than causal tool in this context. The Fine–Gray model should therefore be interpreted as providing a prognostic, population-level benchmark rather than an estimate of causal effect.

A key concern in this study is reverse causation and time-varying confounding. While the within-subject design controls for time-invariant confounders, it cannot adjust for time-varying symptom fluctuations. For example, a relapse may simultaneously trigger both job loss and medication restart, creating a spurious negative association between medication and employment. Similarly, ‘healthy discontinuer’ bias (Sommer et al., 2025) may operate: patients who discontinue medication and remain employed may represent a less severely ill subgroup, inflating apparent negative medication effects. Our lagged exposure sensitivity analyses (Figure 4) partially mitigate this concern, as negative associations persisted across 6, 12, and 24-month lag periods, arguing against reverse causation as the sole explanation. The persistence of effects across progressively longer lag windows suggests a genuine delayed association rather than an artifact of simultaneous relapse and treatment change.

Confounding by indication is partially addressed by the within-subject design, which eliminates between-person severity differences. However, within-person confounding remains possible: periods of medication use may coincide with periods of greater illness severity. The 5–10 year protective effect of medication may partly reflect illness stabilization, treatment optimization, accumulated patient insight into medication management, and stronger therapeutic alliance with clinicians, rather than a purely pharmacological mechanism.

The DREAM employment classification may misclassify ambiguous situations including subsidized employment (fleksjob), parental leave, and informal work arrangements. This nondifferential misclassification would be expected to bias effect estimates toward the null (Copeland et al., 1977; Dosemeci et al., 1990), suggesting our estimates may understate the true magnitude of medication-employment associations.

The F20–F29 diagnostic spectrum encompasses substantial clinical heterogeneity. Initial register-based diagnoses may be revised upon further clinical assessment, introducing potential diagnostic misclassification. Patients who were never medicated during follow-up may represent a less severe subgroup (e.g., individuals with brief psychotic episodes who recovered without pharmacotherapy), and their exclusion from within-subject models limits generalizability to patients with stable medication patterns.

Finally, Denmark’s universal healthcare system, comprehensive social welfare provisions, and active labor market policies may limit generalizability to healthcare systems with less extensive support structures.

### 4.7. Conclusion

Antipsychotic medication is neither purely beneficial nor purely harmful for functional outcomes; its impact evolves with the stage of illness. While standard cohort analyses may suggest a ‘null’ effect, rigorous within-subject modeling reveals a transient trade-off: early-phase treatment serves as a stabiliser that may temporarily constrain vocational capacity before supporting long-term maintenance. Clinical practice should move toward stage-matched rehabilitation, with a specific focus on supporting employment during the ‘consolidation’ years (2–5 post-diagnosis) when the tension between symptom control and functional recovery is most acute.

## 5. Contributions (CRediT)

Conceptualization - RT, ODH, ML, MN, MO Data curation - MO, FHG Formal analysis - RT, MO, CH Methodology - RT, FHG, ODH, ML, MN, MO Software - RT, FHG, MO Supervision - MO Validation - MO, FHG Visualization - RT Writing – original draft - RT Writing – review & editing - RT, FHG, CH, ODH, ML, MN, MO RT, FHG, CH, MN and MO had access to the data.

## 7. Acknowledgements

The authors would like to thank Mathilde Werborg Birkholm and Aleksander Søltoft-Jensen for helping with coding. The authors would also like to thank Janne Petersen for making them part of the team. The authors would like to thank the three anonymous peer reviewers whose comments helped improve this article. The authors would like to express sincere gratitude to each of the 65,630 people whose data were analyzed for this project.

This research was made possible by Danmarks Nationalbank.

## 9. Declaration of Generative AI and AI-assisted technologies in the writing process

During the preparation of this work, the authors used Claude Opus 4.6 (Anthropic, San Francisco, CA) to edit and plan the manuscript. After using this tool, the authors reviewed and edited the content as needed and take full responsibility for the content of the published article.

## Appendix A.

**Supplementary Table 1:** Operationalization of Analytical Categories.

Appendix A. Supplementary Table 1: Operationalization of Analytical Categories
| Category | Label | DREAM Benefit Codes |
| --- | --- | --- |
| *Primary Outcome: Productive Engagement (Categories 3 and 10)* |  |  |
| 10 | Employment | No benefit code (blank/NA indicates labor market attachment) |
| 3 | Education & Vocational Training | 211–219, 231, 232, 299, 412, 413, 511, 521, 522, 651, 652, 661 |
| *Non-Productive States* |  |  |
| 1 | Sickness & Temporary Incapacity | 890–899 |
| 2 | Unemployment & Job-Seeking | 111–113, 115, 121–140, 143–153, 160–169 |
| 4 | Social Assistance & Rehabilitation | 740–781 |
| 5 | Retirement & Pension Benefits | 621, 622, 783, 784, 810, 819, 870, 879, 881, 996, 998 |
| *Censoring Events* |  |  |
| 6 | Emigration | 997 |
| 7 | Death | 999 |

**Figure A.7:**
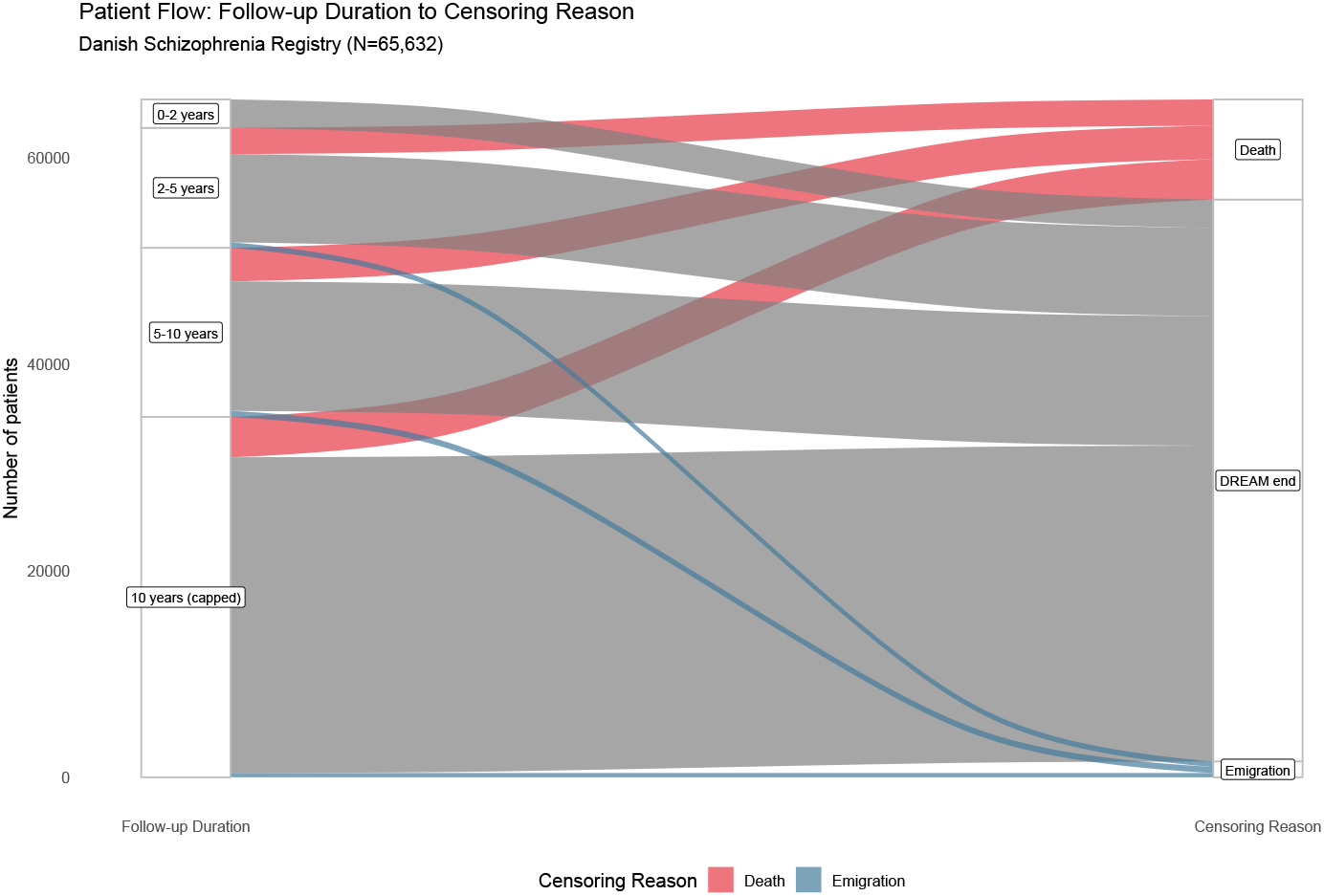
Sankey diagram illustrating censoring flow patterns over 10-year follow-up (n=65,630 patients). Flow width proportional to number of individuals.

**Figure A.8:**
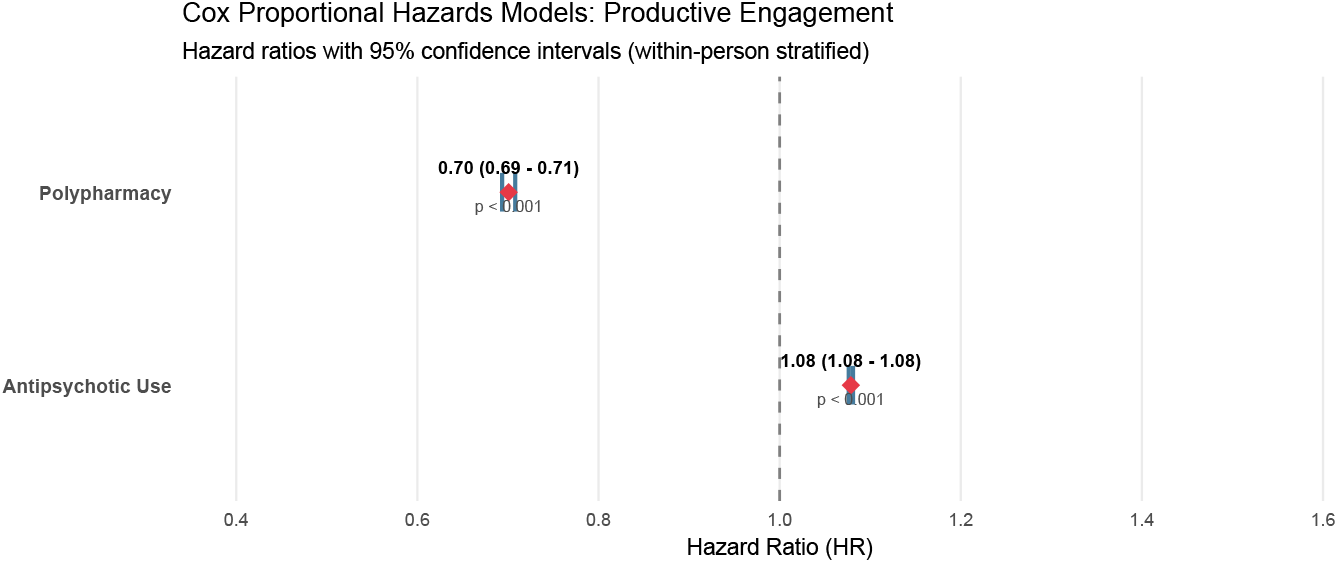
**[Supplementary Figure 1]** Cox proportional hazards forest plot of ‘conditional success rate’ for productive engagement (overall models pooling across time). Hazard ratios with 95% confidence intervals from within-person stratified models (n=50,440 switchers, 21.2 million person-weeks). Model 1: Any antipsychotic use versus none (HR=1.08, p<0.001). Model 2: Polypharmacy versus monotherapy/none (HR=0.70, p<0.001). Dashed vertical line indicates null effect (HR=1.0).

## STROBE Statement: Checklist of Items for Cohort Studies

| Item | Recommendation | Reported | Page |
| --- | --- | --- | --- |
| **Title and Abstract** |  |  |  |
| 1(a) | Indicate study design with commonly used term | “Register-based cohort study” in title and abstract | 1 |
| 1(b) | Provide informative, balanced summary | Abstract provides background, methods, results, interpretation | 1 |
| **Introduction** |  |  |  |
| 2 | Explain scientific background and rationale | Background on antipsychotics, employment, existing evidence gaps | 2 |
| 3 | State specific objectives including prespecified hypotheses | Objectives stated; protocol pre-registered on medRxiv | 2–3 |
| **Methods** |  |  |  |
| 4 | Present key elements of study design | Nationwide cohort, within-subject design, 1998–2023 | 3 |
| 5 | Describe setting, locations, relevant dates | Denmark; diagnosis dates 1998–2023; follow-up to 2023 | 3 |
| 6(a) | Give eligibility criteria, sources/methods of selection | First F20–F29 diagnosis; Danish National Patient Register | 3 |
| 6(b) | For matched studies, give matching criteria and number | Within-subject design; each patient serves as own control | 4 |
| 7 | Define outcomes, exposures, predictors, confounders | Productive engagement (DREAM codes); antipsychotic exposure (N05A); covariates defined | 4–5 |
| 8 | Describe data sources and measurement methods | Danish registers: NPR, DREAM, SMR; ICD-10; ATC codes | 3–4 |
| 9 | Describe efforts to address potential bias | Within-subject design controls time-invariant confounding; competing risks models | 4–5 |
| 10 | Explain how study size was arrived at | All eligible individuals in national registers; N=65,630 | 6 |
| 11 | Explain how quantitative variables were handled | Time-varying binary exposure; person-week observations | 4–5 |
| 12(a) | Describe all statistical methods | Cox models, Fine–Gray, time-stratification, lagged analyses | 4–5 |
| 12(b) | Describe methods for examining subgroups/interactions | Time-stratified by period; medication type subgroups | 5 |
| 12(c) | Explain how missing data were addressed | Complete linkage via CPR; DREAM has no missing benefit data | 3 |
| 12(d) | Describe analytical methods for loss to follow-up | 2-year landmark; death/emigration as competing events | 3, 5 |
| 12(e) | Describe sensitivity analyses | Lagged exposure; SMR integration; time-stratification | 5 |
| **Results** |  |  |  |
| 13(a) | Report numbers at each stage of study | 65,630 included; 1,000 excluded (2-year filter); flow described | 6 |
| 13(b) | Give reasons for non-participation at each stage | Death/emigration within 2 years; see Methods | 3 |

| **Item** | **Recommendation** | **Reported** | **Page** |
| --- | --- | --- | --- |
| 13(c) | Consider use of flow diagram | Sankey diagrams for censoring and medication transitions | Supp. |
| 14(a) | Give characteristics of participants | Table 1: demographics, diagnosis, baseline employment | 6–7 |
| 14(b) | Indicate number with missing data for each variable | Complete data due to mandatory register linkage | 3 |
| 15 | Report numbers of outcome events or summary measures | 26.9 million person-weeks; 48.2% engagement rate; event counts | 6–8 |
| 16(a) | Give unadjusted and adjusted estimates | Time-stratified HRs with CIs; Fine–Gray SHR | 7–10 |
| 16(b) | Report category boundaries for continuous variables | Age groups in Table 1; time periods defined | 6–7 |
| 16(c) | Consider translating relative to absolute risk | Absolute engagement rates by medication type provided | 8 |
| 17 | Report other analyses performed | Lagged effects; SMR sensitivity; dose–response | 9–11 |
| **Discussion** |  |  |  |
| 18 | Summarize key results with reference to objectives | Null Fine–Gray result; temporal heterogeneity in Cox | 12 |
| 19 | Discuss limitations including bias sources | Time-varying confounding; adherence measurement; generalizability | 13 |
| 20 | Give cautious interpretation considering objectives and limitations | Medication as marker vs driver; vocational intervention timing | 12–13 |
| 21 | Discuss generalizability (external validity) | Danish welfare context; universal healthcare; may differ elsewhere | 13 |
| **Other Information** |  |  |  |
| 22 | Give source of funding and role of funders | No direct funding, Danmarks Nationalbank funded research visit; MRC funded salaries; no role in study conduct | 1 |

## References

Ajnakina, O., Stubbs, B., Francis, E., Gaughran, F., David, A., Murray, R., and Lally, J. (2021). Employment and relationship outcomes in first-episode psychosis: A systematic review and meta-analysis of longitudinal studies. Schizophrenia Research, 231:122–133. doi:10.1016/j.schres.2021.03.013

Andersen, M. A., Jensen, T. L., and Petersen, T. S. (2024). Data resource profile: The danish national hospital medicine register. Pharmacoepidemiology and Drug Safety, 33(11):e70054. doi:10.1002/pds.70054

Andersen, P. K., Geskus, R. B., de Witte, T., and Putter, H. (2012). Competing risks in epidemiology: possibilities and pitfalls. International Journal of Epidemiology, 41(3):861–870. doi:10.1093/ije/dyr213

Austin, P. C. and Fine, J. P. (2017). Practical recommendations for reporting fine-gray model analyses for competing risk data. Statistics in Medicine, 36(27):4391–4400. doi:10.1002/sim.7501

Benjamini, Y. and Hochberg, Y. (1995). Controlling the false discovery rate: a practical and powerful approach to multiple testing. Journal of the Royal Statistical Society: Series B (Methodological), 57(1):289–300.

Copeland, K., Checkoway, H., McMichael, A., and Holbrook, R. (1977). The choice of sample size for studies on misclassification. American Journal of Epidemiology, 105(3):239–245. doi:10.1093/oxfordjournals.aje.a112374

Cornblatt, B. A., Carrion, R. E., Addington, J., Seidman, L., Walker, E. F., Cannon, T. D., Cadenhead, K. S., McGlashan, T. H., Perkins, D. O., Tsuang, M. T., Woods, S. W., Heinssen, R., and Lencz, T. (2011). Risk factors for psychosis: Impaired social and role functioning. Schizophrenia Bulletin, 38(6):1247–1257. doi:10.1093/schbul/sbr136

Dosemeci, M., Wacholder, S., and Lubin, J. H. (1990). Does nondifferential misclassification of exposure always bias a true effect toward the null value? American Journal of Epidemiology, 132(4):746–748. doi:10.1093/oxfordjournals.aje.a115716

Fine, J. P. and Gray, R. J. (1999). A proportional hazards model for the subdistribution of a competing risk. Journal of the American Statistical Association, 94(446):496–509. doi:10.1080/01621459.1999.10474144

Hernán, M. A. (2010). The hazards of hazard ratios. Epidemiology, 21(1):13–15. doi:10.1097/EDE.0b013e3181c1ea43

Hjollund, N. H., Larsen, F. B., and Andersen, J. H. (2007). Register-based follow-up of social benefits and other transfer payments: accuracy and degree of completeness in a danish interdepartmental administrative database compared with a population-based survey. Scandinavian Journal of Public Health, 35(5):497–502.doi:10.1080/14034940701271882

Holm, M., Taipale, H., Tanskanen, A., Tiihonen, J., and Mittendorfer-Rutz, E. (2021). Employment among people with schizophrenia or bipolar disorder: A population-based study using nationwide registers. Acta Psychiatrica Scandinavica, 143(1):61–71. doi:10.1111/acps.13254

Jensen, T. B., Andersen, J. T., Jimenez-Solem, E., and Lund, M. (2020). How patients in denmark acquire their medicines: overview, data sources and implications for pharmacoepidemiology. Basic & Clinical Pharmacology & Toxicology, 128(1):46–51. doi:10.1111/bcpt.13472

Kassambara, A., Kosinski, M., and Biecek, P. (2021). survminer: Drawing survival curves using ‘ggplot2’. R package version 0.4.9.

Keepers, G. A., Fochtmann, L. J., Anzia, J. M., Benjamin, S., Lyness, J. M., Mojtabai, R., Servis, M. E., Walaszek, A., Buckley, P., Lenzenweger, M. F., et al. (2020). The american psychiatric association practice guideline for the treatment of patients with schizophrenia. American Journal of Psychiatry, 177(9):868–872. doi:10.1176/appi.ajp.2020.177901

Lau, B., Cole, S. R., and Gange, S. J. (2017). Competing risk regression models for epidemiologic data. American Journal of Epidemiology, 170(2):244–256. doi:10.1093/aje/kwp107

Lawlor, D. A., Tilling, K., and Davey Smith, G. (2017). Triangulation in aetiological epidemiology. International Journal of Epidemiology, 45(6):1866–1886. doi:10.1093/ije/dyw314

Leucht, S., Tardy, M., Komossa, K., Heres, S., Kissling, W., Salanti, G., and Davis, J. M. (2012). Antipsychotic drugs versus placebo for relapse prevention in schizophrenia: a systematic review and meta-analysis. The Lancet, 379(9831):2063–2071. doi:10.1016/S0140-6736(12)60239-6

Lynge, E., Sandegaard, J. L., and Rebolj, M. (2011). The Danish National Patient Register. Scandinavian Journal of Public Health, 39(7_suppl):30–33. doi:10.1177/1403494811401482

Marwaha, S. and Johnson, S. (2004). Schizophrenia and employment. Social Psychiatry and Psychiatric Epidemiology, 39:337–349. doi:10.1007/s00127-004-0762-4

Matsuzaki, H., Hatano, M., Iwata, M., Saito, T., and Yamada, S. (2023). Effectiveness of clozapine on employment outcomes in treatment-resistant schizophrenia: A retrospective bidirectional mirror-image study. Neuropsychiatric Disease and Treatment, 19:615–622. doi:10.2147/NDT.S402945

Moncrieff, J., Crellin, N., Stansfeld, J., Cooper, R., Marston, L., Freemantle, N., Lewis, G., Hunter, R., Johnson, S., Barnes, T., Morant, N., Pinfold, V., Smith, R., Kent, L., Darton, K., Long, M., Horowitz, M., Horne, R., Vickerstaff, V., and Jha, M. (2023). Antipsychotic dose reduction and discontinuation versus maintenance treatment in people with schizophrenia and other recurrent psychotic disorders in england (the radar trial): an open, parallel-group, randomised controlled trial. The Lancet Psychiatry, 10(11):848–859. doi:10.1016/s2215-0366(23)00258-4

Nordfalk, F. and Hoeyer, K. (2017). The rise and fall of an opt-out system. Scandinavian Journal of Public Health, 48(4):400–404. doi:10.1177/1403494817745189

Pedersen, P., Olsen, B. B., Vernal, D. L., Rydborg, M. P., Gasse, C., and Mors, O. (2025). Vocational rehabilitation in young adults with incident schizophrenia—a danish retrospective cohort study. Early Intervention in Psychiatry, 19(6):e70062. e70062 EIP-2024-279.R1. doi:10.1111/eip.70062

Plana-Ripoll, O., Liu, X., Köhler-Forsberg, O., Sørensen, H. T., and Momen, N. C. (2025). Mental disorders in danish hospital registers: A review of content and possibilities for epidemiological research. Clinical Epidemiology, 17:387–407. doi:10.2147/clep.s509147

Pottegård, A., Schmidt, S. A. J., Wallach-Kildemoes, H., Sørensen, H. T., Hallas, J., and Schmidt, M. (2017). Data resource profile: The danish national prescription registry. International Journal of Epidemiology, 46(3):798–798f. doi:10.1093/ije/dyw213

Schoenfeld, D. (1982). Partial residuals for the proportional hazards regression model. Biometrika, 69(1):239–241. doi:10.1093/biomet/69.1.239

Solmi, M., Taipale, H., Holm, M., Tanskanen, A., Mittendorfer-Rutz, E., Correll, C. U., and Tiihonen, J. (2022). Effectiveness of antipsychotic use for reducing risk of work disability: Results from a within-subject analysis of a swedish national cohort of 21,551 patients with first-episode nonaffective psychosis. American Journal of Psychiatry, 179(12):938–946. doi:10.1176/appi.ajp.21121189

Sommer, I. E., de Beer, F., Gangadin, S., de Haan, L., Veling, W., van Beveren, N., Boonstra, N., Rosema, B.-S., van Os, J., Kikkert, M., Koops, S., Noorman, J., Thielen, F., Wijnen, B., Begemann, M., and Consortium, H.-O. (2025). Early dose reduction or discontinuation vs maintenance antipsychotics after first psychotic episode remission: A randomized clinical trial. JAMA Psychiatry. doi:10.1001/jamapsychiatry.2025.2525

Stürup, A. E., Nordentoft, M., Jimenez-Solem, E., Osler, M., Davy, J. W., Christensen, T. N., Speyer, H., Albert, N., and Hjorthøj, C. (2023). Discontinuation of antipsychotics in individuals with first-episode schizophrenia and its association to functional outcomes, hospitalization and death: a register-based nationwide follow-up study. Psychological Medicine, 53(11):5033–5041. doi:10.1017/s0033291722002021

Therneau, T., Crowson, C., and Atkinson, E. (2017). Using Time Dependent Covariates and Time Dependent Coefficients in the Cox Model.

Therneau, T. M. (2024). A package for survival analysis in r. R package version 3.5-8.

Tiihonen, J., Haukka, J., Taylor, M., Haddad, P. M., Patel, M. X., and Korhonen, P. (2011). A nationwide cohort study of oral and depot antipsychotics after first hospitalization for schizophrenia. American Journal of Psychiatry, 168(6):603–609. doi:10.1176/appi.ajp.2011.10081224

Twumasi, R., Lange, M., Hjorthøj, C., Howes, O., Gronemann, F. H., Nordentoft, M., and Osler, M. (2025). Protocol for: Association of antipsychotic medication usage and employment outcomes in patients with schizophrenia spectrum disorders: A danish register-based study. Pre-print. doi:10.1101/2025.10.02.25337161

WHO (2025). Atc classification index with ddds (atc/ddd index 2025). Accessed 2025-10-01. Annual updates are issued each January.

Wolbers, M., Koller, M. T., Witteman, J. C., and Steyerberg, E. W. (2009). Competing risks analyses: objectives and approaches. European Heart Journal, 35(42):2936–2941. doi:10.1093/eurheartj/ehu131

Wunderink, L., Nieboer, R. M., Wiersma, D., Sytema, S., and Nienhuis, F. J. (2013). Recovery in remitted first-episode psychosis at 7 years of follow-up of an early dose reduction/discontinuation or maintenance treatment strategy. JAMA Psychiatry, 70(9):913. doi:10.1001/jamapsychiatry.2013.19

